# Prophylactic post-operative high flow nasal oxygen versus conventional oxygen therapy in obese patients undergoing bariatric surgery (OXYBAR study): a pilot randomised controlled trial

**DOI:** 10.1101/2021.02.03.21251097

**Authors:** Rachel Fulton, Jonathan E Millar, Megan Merza, Helen Johnston, Amanda Corley, Daniel Faulke, Ivan L Rapchuk, Joe Tarpey, Jonathon P Fanning, Philip Lockie, Shirley Lockie, John F Fraser

**Affiliations:** Department of Anaesthesia and Critical Care, Queen Elizabeth University Hospital, Glasgow, United Kingdom; Critical Care Research Group, The Prince Charles Hospital, Brisbane, Australia; Faculty of Medicine, University of Queensland, Brisbane, Australia; Roslin Institute, University of Edinburgh, Edinburgh, United Kingdom; St. Andrew’s War Memorial Hospital, Brisbane, Australia; Griffith University, Griffith, Australia

**Keywords:** Morbid obesity: postoperative complications, postoperative ventilation, morbid obesity and atelectasis

## Abstract

Obesity has become a global pandemic, as a result surgical intervention for weight loss has increased in popularity. Obese patients undergoing operative intervention pose several challenges in respect of their peri-operative care. A prominent feature is the alteration in respiratory mechanics and physiology evident in the obese. These combine to predispose individuals to a reduction in end expiratory lung volume (EELV) and atelectasis after anaesthesia. Consequently, the incidence of post-operative pulmonary complications (PPC) in this cohort has been reported to be in excess of 35%. High flow nasal oxygen (HFNO) has been suggested as a means of increasing EELV in post-operative patients, reducing the likelihood of PPC. We conducted a single centre, pilot, randomised controlled trial (RCT) of conventional oxygen therapy versus HFNO in patients after bariatric surgery. The aim of the study was to investigate the feasibility of using Electrical Impedance Tomography (EIT) as a means of assessing respiratory mechanics and to inform the design of larger, definitive RCT. Fifty patients were randomised during a 10-month period (conventional O_2_ n=25 vs. HFNO n = 25). One patient crossed over from conventional O_2_ to HFNO. There was no loss to follow-up. and analyses were performed on an intention-to-treat basis. Delta EELI was higher at 1 hour in patients receiving HFNO (mean difference = 831 Au (95% CI -1636 – 3298), p = 0.5). Continuous EIT beyond 1 hour was poorly tolerated. At 6 hours, there were no differences in PaO_2_/FiO_2_ ratio or PaCO_2_. ICU and hospital LOS were comparable. Only one patient developed a PPC (in the HFNO group). In a secondary analysis, delta EELI was positively correlated with increasing BMI. These data suggest that a large-scale randomised controlled trial of HFNO after bariatric surgery in an ‘all-comers’ population is likely infeasible. Furthermore, while EIT is a useful tool for assessing respiratory mechanics in this group it could not be considered a patient-centred outcome in a larger study. Similarly, the infrequency of PPC precludes its use as a primary outcome in a definitive trial. Future studies should focus on identifying patients most at risk for post-operative pulmonary complications and those in whom HFNO is likely to confer greatest benefit.

## Introduction

Obesity is a global pandemic, with over 2 billion individuals now classed as overweight or obese [1]. As a result, the use of surgical intervention in the management of obesity has increased in popularity [2]. Surgery, in selected individuals, has been shown to be a safe and effective means of achieving sustained weight loss [3]. Despite the success of operative interventions, patients undergoing these procedures continue to represent a challenge in respect of their peri-operative and anaesthetic management.

Obesity is associated with a higher mortality after non-cardiac surgery and with an increased risk of developing a variety of post-operative complications, including myocardial infarction and wound infection [4]. The adverse impact of obesity on respiratory mechanics and ventilation is also well described. Obesity tends to increase the work of breathing and promote rapid, shallow breathing. In turn, end expiratory lung volume (EELV) is reduced, disposing the individual toward atelectasis and ventilation/perfusion mismatching [5-8]. In those undergoing bariatric surgery, the prevalence of post-operative atelectasis has been reported to be as high as 38% [9]. However, the ability to directly translate these physiological alterations into observable morbidity and mortality remains uncertain [10-12].

The post-operative application of high flow nasal oxygen (HFNO) has been suggested as a means of reducing the risk of pulmonary complications [13]. Previous studies have reported the physiological benefits of HFNO in the post-operative period, associating its use with improvements in EELV and gas exchange [14].

We conducted a pilot, randomised controlled trial of HFNO versus conventional oxygen therapy after bariatric surgery to investigate the physiological action of HFNO in the obese, assess the feasibility of using an Electrical impedance Tomography (EIT) measure of EELV as an outcome, and to inform the design of a larger, definitive trial.

## Methods

This single centre study was approved by the UnitingCare Health and Human Research Ethics Committee (UCH/HERC/1709). The trial was prospectively registered with the Australian New Zealand Clinical Trials Registry (ACTRN12617000694314). Prospective written informed consent was obtained from all patients prior to their inclusion in the study. This study has been reported in compliance with the CONSORT extension for pilot randomised and feasibility trials [15].

### Study setting

St. Andrew’s War Memorial Hospital (SAWMH), Brisbane, is a tertiary referral centre with a 15-bed general adult ICU. Bariatric surgical services are provided by a single specialist surgeon, conducting over 250 procedures per year, including; laparoscopic gastric bypass, sleeve gastrectomy, and gastric banding. All patients scheduled to undergo laparoscopic surgical intervention for weight loss were screened for eligibility between April 2017 and January 2018. The trial was scheduled to finish after the inclusion of 50 patients. This number was based on an audit of surgical workload over a 12-month period, with an assumed randomisation to screening ratio of 1:2 (10% loss to follow-up) and a planned study period of 14 months.

### Inclusion, exclusion, and randomisation

Patients were eligible for inclusion in the study if they were aged ≥ 18 years and had a body mass index (BMI) ≥ 30 kg/m^2^. We excluded patients if they had a contraindication to the use of HFNO, had a chest circumference too large to allow for the use of an Electrical Impedance Tomography (EIT) belt (> 150 cm), or if they had evidence of pre-operative hypoxaemia (SpO_2_ < 92% on air) and/or severe chronic lung disease.

Prior to surgery, patients were randomised in a 1:1 fashion (using a computer-generated sequence based on block sizes of 4, 6, or 8). An investigator, not involved in the peri-operative care of patient, obtained the allocation in a sealed envelope and informed the Intensive Care Unit staff. The surgical and anaesthetic teams were blinded to the patient’s allocation until return to the ICU, after which treatment allocation was unblinded.

### Study protocol

The study protocol was pre-defined and has been published online [16]. In brief, the conduct of general anaesthesia was at the discretion of the responsible anaesthetist. Given the nature of the surgery, all patients underwent endotracheal intubation and mechanical ventilation. All patients were extubated in theatre and received supplemental oxygen via a Hudson mask at 6 L/min before immediate transfer to the ICU. Those randomised to the conventional oxygen group continued to receive supplemental oxygen via a face mask, titrated to achieve a peripheral oxygen saturation (SpO_2_) ≥ 95%. If the patient consistently achieved or exceeded this target, the level of support was reduced in stages to a minimum of standard nasal cannula providing 2 L/min oxygen. Patients randomised to the intervention group received supplemental oxygen via high flow nasal cannulae (Airvo™ 2, Fisher & Paykel, New Zealand). HFNO was commenced at a flow rate of 50 L/min and set to provide an inspired fraction of oxygen (FiO_2_) of 0.5. Inspired oxygen concentrations were titrated to maintain a SpO_2_ ≥ 95%. A constant flow was maintained for the duration of the study. Oxygen therapy was protocolised for 6 hours after which care reverted to the discretion of the treating intensivist. HFNO could be continued if deemed desirable by the clinical team. Patients were discharged from the ICU on post-operative day one, unless a higher level of care was judged to be required by the intensive care team. Timing of hospital discharge was decided upon by the admitting surgeon.

### Outcome assessment

The primary outcome of the study was change in end expiratory lung impedance (ΔEELI) at 60 minutes, measured using EIT (Pulmovista 500, Dräger, Lübeck, Germany). EIT is a non-invasive, bedside imaging technique used to acquire data on regional lung ventilation and volume. It’s use as a tool for investigating patients receiving HFNO is well validated [14, 17, 18]. Previous studies have shown that changes in End Expiratory Lung Impedance (EELI), measured using EIT are positively correlated with changes in EELV [14]. Likewise, tidal variation, measured by EIT, is related to changes in tidal volumes [14].

*A priori*, we defined ΔEELI at 6 hours as the primary outcome measure [16]. During the trial it became apparent that the continuous placement of an EIT belt for > 2-3 hours was intolerable for most patients. Removal and re-application of the belt invalidates the measurement of EELI and thus 60 minutes was adopted as a primary endpoint prior to the completion of the trial and analysis of the data. EIT analysis was conducted using the manufacturers software (EIT Data Analysis Tool v6.1, Dräger, Lübeck, Germany) according to our previously published methods [14]. Pre-defined secondary outcomes were: ΔEELI at 15 and 30 minutes; tidal variation at 15, 30 and 60 minutes (measured by EIT); change in PaO_2_/FiO_2_ ratio, PaCO_2_ and respiratory rate (RR) at 60 minutes, 3 and 6 hours; change in modified Borg dyspnoea score [19] at 60 minutes, 3 and 6 hours; simple numeric pain reporting scale at 60 minutes, 3, and 6 hours; requirement for escalation of oxygen therapy; ICU length of stay; re-admission to the ICU; hospital length of stay; and hospital re-admission in the first 6 weeks. In addition, patients were followed up until 6 weeks post-operatively to identify those with a diagnosis of a new post-operative pulmonary complication (PPC), defined as any pulmonary abnormality, disease, or dysfunction that adversely affected the clinical course of the patient.

### Statistical analysis

All analyses were on an intention-to-treat basis and performed with GraphPad Prism v8.0.1 (GraphPad Software, San Diego, CA, USA). Data are expressed as mean (± SD) if normally distributed or median (IQR) if non-normally distributed. Longitudinal data were analysed by fitting a mixed model. This mixed model used a compound symmetry covariance matrix and was fit using Restricted Maximum Likelihood (RML). Where a significant interaction was observed between groups or between time and group, post-hoc comparisons were undertaken using Fisher’s Least Significant Difference (LSD) test. Alternatively, non-longitudinal data were compared using an unpaired t-test if normally distributed, or a Mann-Whitney test if non-normally distributed. Categorical data were compared using the chi-squared test. Adjustment for multiple comparisons was made using a False Discovery Rate (FDR) method (Benjamini-Hochberg). The FDR was restricted to 5%. Correlation between variables was assessed using Pearson’s correlation co-efficient. A *P* value < 0.05 was considered significant.

## Results

Fifty-six patients were screened for enrolment, of whom 50 were randomised (25 patients per group) (Fig. 1). All patients completed the study protocol. One patient randomised to receive conventional oxygen was escalated to HFNO at hour 4 due to hypoxaemia. There was no loss to follow-up. Baseline characteristics and intra-operative parameters are detailed in Table 1. There were no adverse events related to either HFNO or EIT use.

**Table 1.** Patient characteristics at baseline. BMI – body mass index; CPAP – continuous positive airway pressure; COPD – chronic obstructive pulmonary disease; NMB – neuromuscular blocking drug; OSA – obstructive sleep apnoea.

|  | **Conventional O<sub>2</sub>**<br>(n=25) | **HFNO**<br>(n=25) |
| --- | --- | --- |
| **Age** (years), mean (±SD) | 46 ± 9.2 | 48 ± 11.1 |
| **Female** (n (%)) | 20 (80) | 19 (76) |
| **BMI** (kg/m<sup>2</sup>), mean (±SD) | 44.4 ± 6.9 | 43.1 ± 7.4 |
| > 40 (n (%)) | 18 (72) | 14 (56) |
| > 50 (n (%)) | 9 (36) | 5 (20) |
| **Prior respiratory disease** (n (%)) |  |  |
| Asthma | 4 (16) | 4 (16) |
| OSA | 14 (56) | 9 (36) |
| Using CPAP | 14 (56) | 6 (24) |
| COPD | 0 | 0 |
| Other | 2 (8) | 0 |
| **Prior cardiac disease** (n (%)) |  |  |
| Hypertension | 7 (28) | 9 (36) |
| Ischaemic heart disease | 1 (4) | 0 |
| Other | 3 (12) | 4 (16) |
| **Smoking status** (n (%)) |  |  |
| Never smoked | 9 (36) | 14 (56) |
| Ex-smoker | 15 (60) | 10 (40) |
| Current smoker | 1 (4) | 1 (4) |
| **Type of surgery** (n (%)) |  |  |
| Gastric band | 2 (8) | 3 (12) |
| Sleeve gastrectomy | 16 (64) | 14 (56) |
| Gastric bypass | 4 (16) | 3 (12) |
| Other | 3 (12) | 5 (20) |
| **Operative duration** (mins), mean (±SD) | 142 ± 32 | 137 ± 40 |
| **Intra-operative crystalloid volume** (mL), mean (SD) | 1444 ± 381 | 1428 ± 403 |
| **Sugammadex NMB reversal** (n (%)) | 19 (76) | 20 (80) |

**Figure 1.**
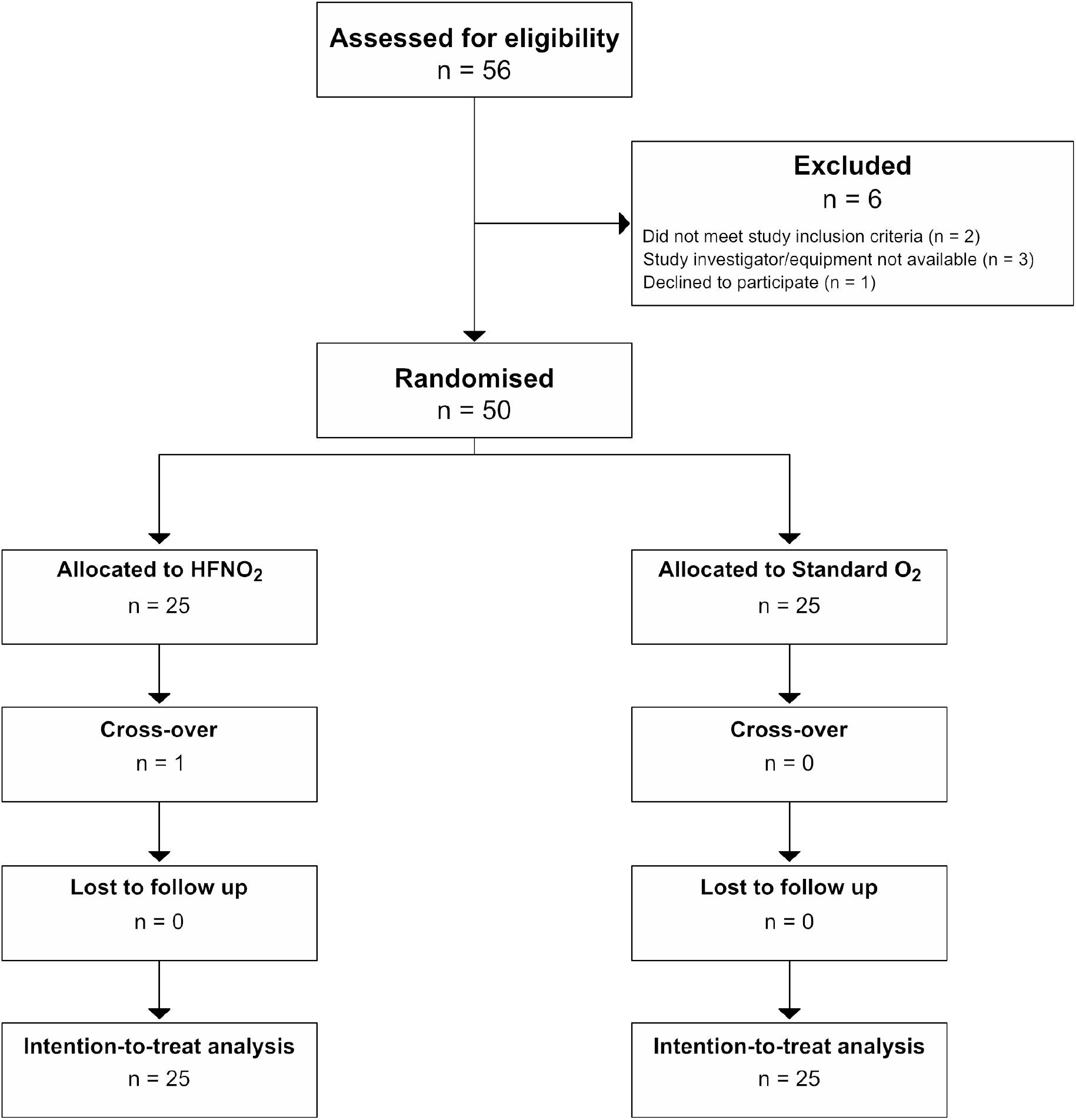
CONSORT flow chart for the study.

### End expiratory lung impedance and tidal variation

Overall, 46 (92%) patients had ≥ 3 analysable EIT recordings (conventional O_2_ n = 23 vs. HFNO n = 23). One patient in each group did not have EIT data fit for analysis due to corruption of the baseline recording. There were no significant differences in change in EELI from baseline (ΔEELI) between groups (Fig. 2). However, the group receiving HFNO showed a consistent increase in EELI over 60 minutes, while the group receiving conventional oxygen therapy had a diminishing improvement over the first 30 minutes and failed to match the improvement seen at baseline by 60 minutes (HFNO vs. conventional O_2_ mean difference at 60 minutes = 831 Au (95% CI -1636 – 3298), p.adj = 0.5). There were no significant differences in tidal variation between groups (Fig. 2).

**Figure 2.**
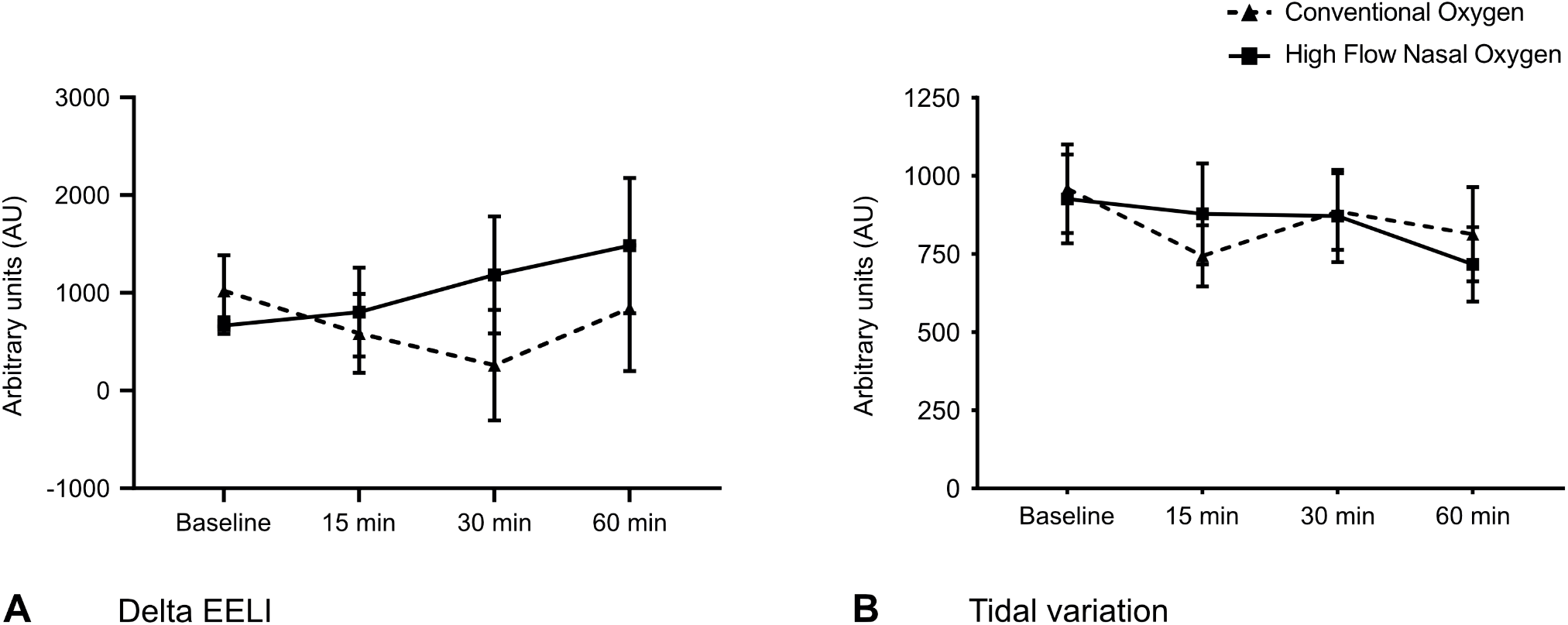
Electrical impedance tomography parameters. A. Delta End Expiratory Lung Impedance (ΔEELI). B. Tidal variation. Data are expressed as mean (± 95% confidence interval).

### Oxygenation and gas exchange

Oxygenation, as measured by PaO_2_/FiO_2_ ratio, was similar between groups throughout the study (Fig. 3). This was despite the conventional oxygen group receiving a significantly lower FiO_2_ from 1 hour onwards. Both groups exhibited a progressive reduction in PaCO_2_ during the study period (Fig. 3).

**Figure 3.**
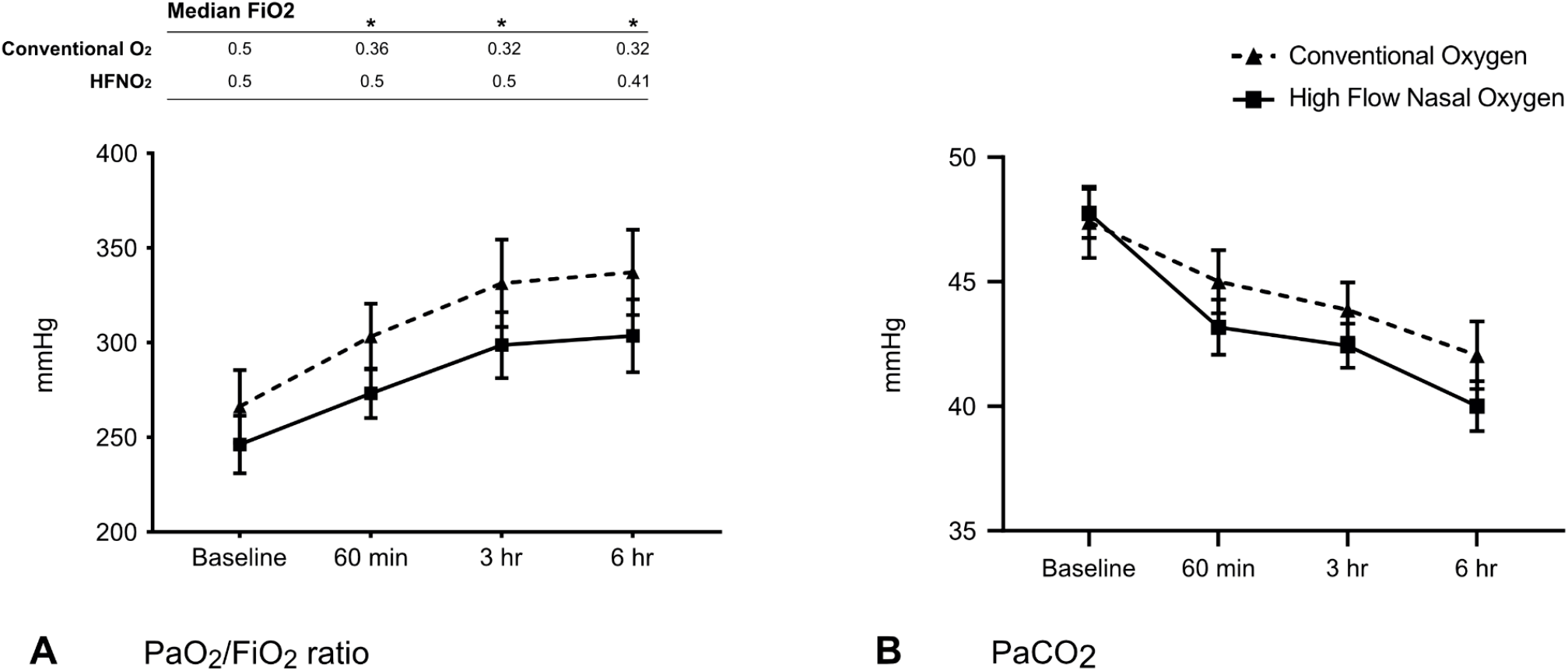
Gas exchange parameters. A. Arterial partial pressure of oxygen to inspired fraction of oxygen ratio (PaO_2_/FiO_2_) and median FiO_2_. B. Arterial partial pressure of carbon dioxide (PaCO_2_). Data are presented as mean (±95% confidence interval). * p < 0.01.

### Dyspnoea and pain scores

Patient reports of dyspnoea, measured using a modified Borg score, were universally low. There were no differences observed between groups (Table 2). Pain scores varied across time points and were higher in those receiving HFNO at 6 hours (Table 2).

**Table 2.** Dyspnoea scores, pain scores, and patient outcomes. ICU – intensive care unit; LOS – length of stay; PPC – post-operative pulmonary complication.

|  | **Conventional O<sub>2</sub>**<br>(n=25) | **HFNO**<br>(n=25) | **P value** |
| --- | --- | --- | --- |
| **Borg Dyspnoea Score [0-10] (median (IQR))** |  |  |  |
| Baseline | 0 (0-0) | 0 (0-0) | 0.387 |
| 1 hour | 0 (0-0) | 0 (0-0) | 0.322 |
| 3 hours | 0 (0-0) | 0 (0-0) | 0.389 |
| 6 hours | 0 (0-0) | 0 (0-0) | 0.180 |
| **Pain Score [0-10] (median (IQR))** |  |  |  |
| Baseline | 3 (2-6) | 1.5 (0-4.25) | 0.107 |
| 1 hour | 3 (0-5) | 3.5 (0-6.25) | 0.397 |
| 3 hours | 3 (1-4) | 2.35 (0-3.25) | 0.691 |
| 6 hours | 0 (0-3) | 2.79 (1.5-4.5) | 0.026 |
| **ICU LOS (days)** | 1 (1-1) | 1 (1-1) | 0.982 |
| **ICU re-admission (n (%))** | 1 (4) | 0 | 1 |
| **Hospital LOS (days)** | 2 (2-2) | 2 (2-2) | 0.617 |
| **Hospital re-admission (n (%))** | 0 | 4 (16) | 0.128 |
| **PPC in first 6 weeks (n (%))** | 0 | 1 (4) | 0.128 |

### Length of stay and post-operative complications

Intensive care unit and hospital length of stay (LOS) were equivalent between groups. One patient in the conventional O_2_ group required readmission to ICU after revisional surgery for an anastomotic leak. Only one patient (from the HFNO group) was diagnosed with a PPC during the 6-week follow-up period, in this case atelectasis with small bilateral pleural effusions. Four patients in the HFNO required readmission to hospital following their initial discharge, none of these incidents were related to pulmonary complications.

### Association between BMI and ΔEELI

The association between BMI and ΔEELI was assessed in a post-hoc analysis. There was no significant correlation between BMI and ΔEELI at any time point in the conventional group. However, in those receiving HFNO, increasing BMI was positively correlated with higher ΔEELI values at 30 and 60 minutes (Fig. 4).

**Figure 4.**
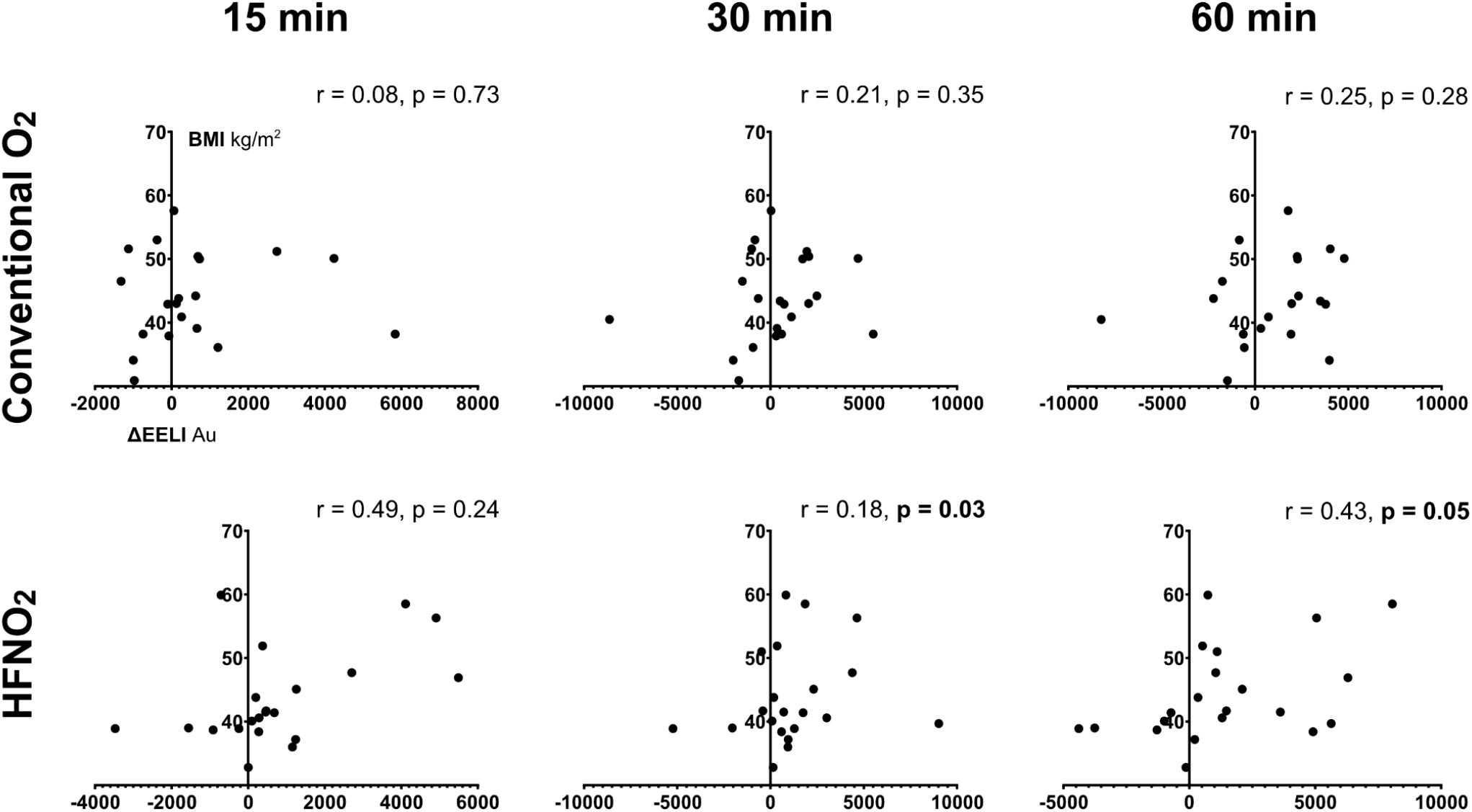
Association between BMI and ΔEELI. Au – arbitrary units.

## Discussion

We conducted a single centre, pilot, randomised controlled trial of HFNO versus conventional oxygen therapy, in patients undergoing bariatric surgery. Our principal findings can be summarised as; (1), HFNO failed to produce a significant increase in EELI at 60 minutes, (2), similarly, the use of HFNO was not associated with improvements in PaO_2_/FiO_2_ ratio, dyspnoea, length of stay, or the incidence of post-operative pulmonary complications, (3), the use of HFNO was associated with higher pain scores at 6 hours, and (4), the effect of HFNO on EELI was enhanced in patients with a higher BMI. EIT was a feasible method of assessing respiratory mechanics in this population, however, in a definitive trial, would not constitute a patient-centred outcome and is not suitable for use over extended periods.

Additionally, the low incidence of post-operative pulmonary complication in our study would suggest that its use as a primary outcome would likely require an infeasibly large sample size. Future effectiveness studies in this area may consider enrichment by including patients with a higher BMI or those with other risk factors for post-operative pulmonary complication.

The ability of HFNO to improve EELI post-operatively has been studied previously [14, 20]. In a cohort of patients undergoing elective cardiac surgery, Corley et al. described an improvement in EELI in patients transitioning from low flow oxygen therapy to HFNO. The magnitude of this effect was positively correlated with BMI. These findings are consistent with the relationship between EELI and BMI in patients randomised to HFNO in our study. In contrast, Corley et al. did report reductions in dyspnoea and tidal variation. In a mixed critical care population, Zhang et al. evaluated the physiological effects of HFNO following extubation [20]. This study demonstrated the ability of HFNO to increase EELI and reduce atelectasis. However, this was variable, with several patients impervious to recruitment and a minority exhibiting evidence of regional over distension despite an increase in global EELI. There was no difference in BMI between the so-called ‘high’ and ‘low’ potential for recruitment groups. Similar to our findings, Zhang et al. failed to record a change in tidal variation.

In patients randomised to HFNO in our study, pain scores were higher at 6 hours. The high gas flow rates used during HFNO have been associated with increased abdominal distension [21], of particular concern after gastric surgery and a possible explanation for our findings. An alternative explanation may be the nasal discomfort reported by some patients receiving HFNO [22].

The incidence of post-operative pulmonary complications in our study was 2% across both groups. This is much lower than the 25.7% seen in conventional oxygen therapy or 27% in HFNO, described in a recent meta-analyses of trials comparing the two [23]. However, the majority of studies included in this review enrolled patients after cardiac or thoracic surgery. Our findings are more consistent with the incidence of PPC after non-cardiac surgery [24] and in particular with the very low incidence (<1%) of PPC in a large series of bariatric surgical patients [25].

This study has several limitations. First, we were unable to perform analyses of EIT data beyond 60 minutes. While this was an important finding from the perspective of feasibility, it may be that prolonged observation would have demonstrated a continued and significant divergence in EELI between groups. Second, we chose to start the respiratory support protocol on return to the ICU. This may have permitted decruitment in the period between extubation and commencing therapy, which may have been overcome by choosing to extubate on to HFNO. However, this was a pragmatic decision given the difficulty of transferring patients on HFNO with current technology. Third, this was a single centre study, with patients electively admitted to a critical care bed for at least 24 hours post-operatively, this may not be generalisable to practice elsewhere and may have contributed to the low incidence of post-operative pulmonary complication. Finally, this was a pilot study and was not powered to detect differences in EIT or clinical outcomes and should be interpreted in that light.

In summary, our findings suggest that a large-scale randomised controlled trial of HFNO after bariatric surgery in an ‘all-comers’ population is likely infeasible. Future studies should focus on identifying patients most at risk for post-operative pulmonary complications and those in whom HFNO is likely to confer greatest benefit.

## Data Availability

Source trial data will be made available upon reasonable request.

## Notes

### Competing Interest Statement

JEM has received honoraria for educational activity from Fisher and Paykel. JFF has received research support from Fisher and Paykel. Fisher and Paykel were not involved in the funding, design, conduct, or interpretation of this study.

### Clinical Trial

ACTRN12617000694314

### Clinical Protocols

https://trialsjournal.biomedcentral.com/articles/10.1186/s13063-018-2777-2

### Funding Statement

The study, including research nursing infrastructure, was supported by a grant from Wesley Medical Research (https://www.wesleyresearch.org.au).

### Author Declarations

This single centre study was approved by the UnitingCare Health and Human Research Ethics Committee (UCH/HERC/1709).

